# First wastewater surveillance-based city zonation for effective COVID-19 pandemic preparedness powered by early warning: A study of Ahmedabad, India

**DOI:** 10.1101/2021.03.18.21253898

**Authors:** Manish Kumar, Madhvi Joshi, Anil V. Shah, Vaibhav Srivastava, Shyamnarayan Dave

## Abstract

Following the proven concept, capabilities, and limitations of detecting the RNA of Severe Acute Respiratory Coronavirus 2 (SARS-CoV-2) in wastewater, it is pertinent to understand the utility of wastewater surveillance data on various scale. In the present work, we put forward the first wastewater surveillance-based city zonation for effective COVID-19 pandemic preparedness. A three-month data of Surveillance of Wastewater for Early Epidemic Prediction (SWEEP) was generated for the world heritage city of Ahmedabad, Gujarat, India. In this expedition, one hundred sixteen wastewater samples were analyzed to detect SARS-CoV-2 RNA, from September 3^rd^ to November 26^th^, 2020. A total of 111 samples were detected with at least two out of three SARS-CoV-2 genes (N, ORF 1ab, and S). Monthly variation depicted a significant decline in all three gene copies in October compared to September 2020, followed by a sharp increment in November 2020. Correspondingly, the descending order of average genome concentration was: November (∼10729 copies/ L) > September (∼3047 copies/ L) > October (∼454 copies/ L). Monthly variation of SARS-CoV-2 RNA in the wastewater samples may be ascribed to a decline of 19.3% in the total number of active cases in October 2020 and a rise of 1.82% in November 2020. Also, the monthly recovery rate of patients was 16.61, 19.31, and 15.58% in September, October, and November 2020, respectively. The percentage change in the genome concentration was observed in the lead of 1-2 weeks with respect to the provisional figures of confirmed cases. SWEEP data-based city zonation was matched with the heat map of the overall COVID-19 infected population in Ahmedabad city, and month-wise effective RNA concentration variations are shown on the map. The results expound on the potential of WBE surveillance of COVID-19 as a city zonation tool that can be meaningfully interpreted, predicted, and propagated for community preparedness through advance identification of COVID-19 hotspots within a given city.

**Graphical Abstract:** 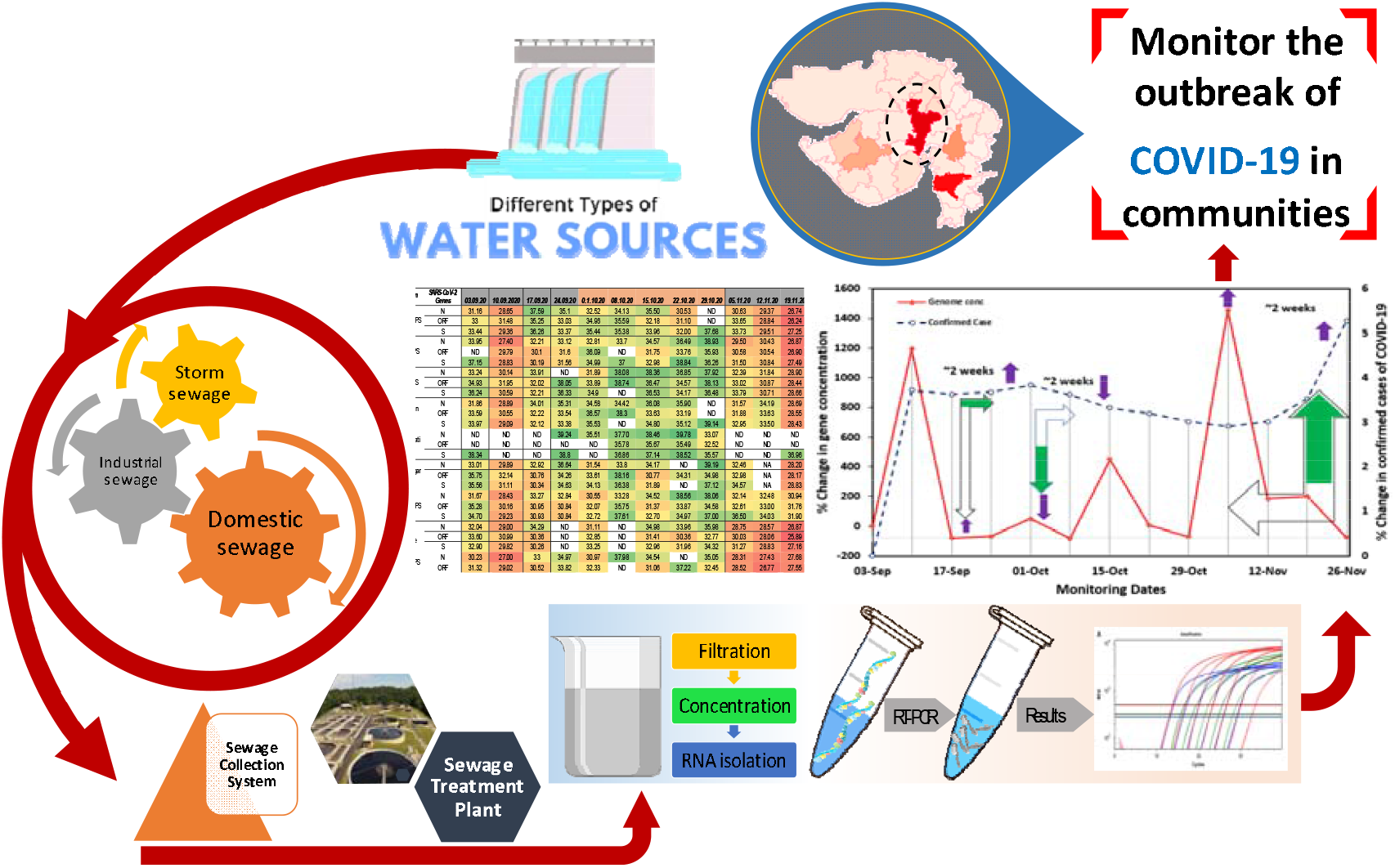

**Highlights:**

▪ Wastewater surveillance-based city zonation is effective for COVID-19 pandemic preparedness.
▪ Three months variation of SARS-CoV-2 RNA in the wastewaters of Ahmedabad, India is presented.
▪ Wastewater monitoring offers a lead of ^∼^2 weeks to realize and manage the pandemic situation.
▪ Mapping powered by early warning can strengthen the preparedness of community.
▪ WBE based COVID-19 surveillance is a high end technique for identifying hotspots on city scale.

## 1. Introduction

The contagious severe acute respiratory syndrome coronavirus 2 (SARS-CoV-2), responsible for the coronavirus pandemic, has infected 11 million people in India alone by February 22^nd^, 2021 (WHO, 2020). A large number of asymptomatic patients exerted a never seen before challenges over the actual estimation of disease spread based on clinical surveillance (Rimoldi et al., 2020; Medema et al., 2020). Earlier studies suggested that 18-45% of patients do not have signs of infection with COVID-19 but are capable of spreading the disease and pose an adverse impact on the actual containment of the disease (Lavezzo et al., 2020; Yang et al., 2020; Mizumoto et al., 2020; Nishiura et al., 2020). As upto 67% of infected people showed SARS-CoV-2 presence in feces (Chan et al., 2020; Cheung et al., 2020; Parasa et al., 2020; Wong et al., 2020), alternative approaches such as wastewater-based epidemiology (WBE) surveillance has gained loads of recognition as a viable option that can provide early warning of the upcoming prevalence of the disease within a community (Tran et al., 2020; Hata et al., 2021; Kumar et al., 2021a, b,). One of the advantages of WBE is that wastewater contains feces from a huge number of people. Therefore, it may require a far fewer number samples and less labor than clinical testing to know the presence of infected persons in the area (Prevost et al., 2015). However, the sensitivity of WBE for SARS-CoV-2 detection is comparatively less than norovirus, presumably due to the low SARS-CoV-2 load in the patient’s fecal matter and it’s enveloped nature (Hata et al., 2020). Also, to evaluate WBE’s potential as an early prediction tool for COVID-19 pandemic, it is essential to explore the correlation between the SARS-CoV-2 genetic load in wastewater and the number of cases at the district level in each country.

Overall, following the proven concept and capabilities of detecting the RNA of Severe Acute Respiratory Coronavirus 2 (SARS-CoV-2) in wastewater, several limitations and bottlenecks have been put forward towards its practical applicability (Zhu et al., 2021). On the other hand, there is a dire need for time-series data of SARS-CoV-2 RNA concentration in the wastewater that can be matched with the actual clinical survey data to confirm the utility and predictability of wastewater surveillance. This is also imperative for the adaptation of the *Surveillance of Wastewater for Early Epidemic Prediction (SWEEP)* on the policy level, which has been for some reason still delayed in the major parts of the globe (<u>Tiwari et al.,2021</u>). There has also been an active debate of varying levels of effectiveness of WBE based on the size of watersheds, catchment type, complexity of sewer systems, and population. Although the science, concepts, and knowledge pertaining to COVID-19 are still evolving and changes rapidly, it is pertinent to check how effective SWEEP can be on the urban scale, that too if cases reported from the given city have been pretty high. Under this scenario, the four major directions in the field of SWEEP may be summarised as i) substantiating the data unravelling the early warning capability of wastewater surveillance for COVID-19 through temporal studies on SARS-CoV-RNA detection; ii) need for the escalation of WBE monitoring of various parts of the globe to generate results from all the levels of COVID-19 situation; iii) developing the model that can use Ct-value obtained through SWEEP into the meaningful predictions for effective COVID-19 pandemic preparedness; and iv) collectively reach to the understanding of critical issues like removal, discharge, decay, dilution, and infectivity due to the presence of SARS-CoV-2 RNA in wastewaters (Kumar et al., 2021).

In view of this, the objective of this study was to put forward the evidence of practical applicability of SWEEP for COVID-19 pandemic management by comparing the detected concentration of SARS-CoV-2 RNA in wastewater of various parts of the city with the COVID-19 clinical cases. The Idea is that the clinical surveillance hardly classify the city into precise zones where more tests or attention are required, while SWEEP-based information can help zoning of the city and identifying the hotspots on a city scale. The detected concentrations of SARS-CoV-2 RNA in wastewater would reflect the true prevalence of COVID-19 infection in the sewer catchment, including clinically undiagnosed patients, while the number of clinically reported cases covers only diagnosed patients and also depends on the number of PCR diagnosis. We analyzed SARS-CoV-2 RNA in the wastewater samples (n = 116) from nine different locations, including wastewater pumping stations and sewage treatment plant (STP) and in Ahmedabad, India, from September 3^rd^ to November 26^th^, 2020 (thirteen weeks), with the following objectives: a) detection and quantification of SARS-CoV-2-RNA concentration in the influent wastewater samples of Ahmedabad to understand the temporal variation in the pandemic situation over three months, b) weekly resolution of the SARS-CoV-2 RNA data for three months in wastewater samples; and c) explicating the potential of WBE for COVID-19 surveillance as a potential tool for identifying hotspots and public health monitoring at the city level.

## 2. Material and Methods

### 2.1 Sampling approach

Wastewater samples were collected from nine different locations, including eight wastewater pumping stations and a single sewage treatment plant (**Fig. 1**). The samples were collected weekly for thirteen weeks from each location during September to November 2020. A total of 116 samples were analyzed in the present study to detect SARS-CoV-2 RNA from nine different sites, comprising 103 samples from eight wastewater pumping stations and 13 samples from a single sewage treatment plant in Ahmedabad, India. All the samples were collected by grab hand sampling using 250 ml sterile bottles (Tarsons, PP Autoclavable, Wide Mouth Bottle, Cat No. 582240, India). Simultaneously, blanks in the same type of bottle were examined to know any contamination during the transport. The samples were kept cool in an ice-box until further process. The analysis was performed on the same day after bringing the samples to the laboratory. All the analyses were performed in Gujarat Biotechnology Research Centre (GBRC), a laboratory approved by the Indian Council of Medical Research (ICMR), New Delhi.

**Fig. 1.**
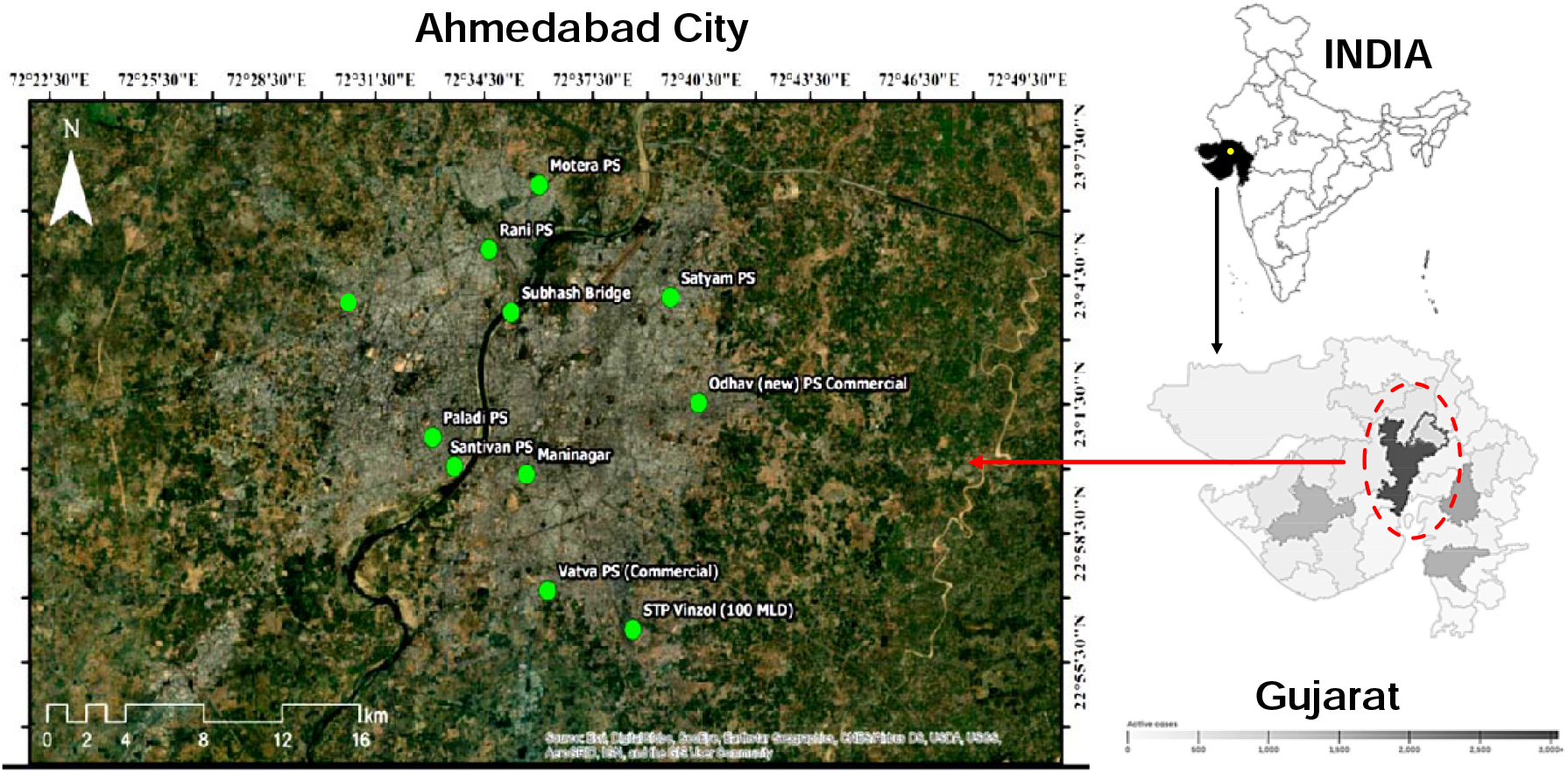
Geospatial position of sampling locations in Ahmedabad city

### 2.2 Detection and extraction of viral RNA from wastewater samples

#### 2.2.1 Precipitation of viral particle

30 mL samples were centrifuged at 4000×g (Model: Sorvall ST 40R, Thermo Scientific) for 40 minutes in a 50 mL falcon tube followed by filtration of supernatant using 0.22-micron syringe filter (Mixed cellulose esters syringe filter, Himedia). After filtrating 25 mL of the supernatant, 2 g of PEG 9000 and 0.437 g of NaCl (17.5 g/L) were mixed in the filtrate, and this was incubated at 17°C, 100 rpm overnight (Model: Incu-Shaker™ 10LR, Benchmark). Next day, the mixture was centrifuged at 14000×g (Model: Kubota 6500, Kubota Corporation) for about 90 minutes. The supernatant was discarded after centrifugation, and the pellet was resuspended in 300µL RNase free water. The concentrated sample was kept in 1.5ml eppendorf at −40 °C, and this was further used as a sample for RNA isolation.

#### 2.2.2 RNA isolation, RT-PCR and gene copy estimation

RNA isolation from the pellet with the concentrated virus was performed using NucleoSpin® RNA Virus isolation kit (Macherey-Nagel GmbH & Co. KG, Germany). The samples were spiked with MS2 phage as an internal control prior to the RNA extraction provided by TaqPathTM Covid-19 RT-PCR Kit. Some other specifics are, a) the nucleic acid was extracted by NucleoSpin® RNA Virus isolation kit and Qubit 4 Fluorometer (Invitrogen) was used for the total RNA concentrations estimation, b) MS2 phage was taken as a molecular process inhibition control for evaluating the efficiency of nucleic acid extraction and PCR inhibition. (MPC; Haramoto et al., 2018). Briefly, steps were carried out as per the guideline provided with the product manual of Macherey-Nagel GmbH & Co. KG, and RNAs were detected using real-time PCR (RT-PCR).

Applied Biosystems 7500 Fast Dx Real-Time PCR Instrument (version 2.19 software) was used for SARS-CoV-2 gene detection. In the process, the probes anneal to three specific target sequences located between three unique forward and reverse primers for the N, ORF 1ab, and S genes. A template of 7 µl of extracted RNA was used in each reaction with TaqPath™ 1 Step Multiplex Master Mix (Thermofischer Scientific, USA). Total reaction mixture volume of 20 µL contained 10.50 µL Nuclease-free Water, 6.25 µL Master Mix, and 1.25 µL COVID-19 Real-Time PCR Assay Multiplex. Three controls were used, namely: positive control (TaqPath™ COVID 19 Control), one negative control (from extraction run spiked with MS2), and no template control (NTC). The real-time PCR contained 1 incubation step cycle of 25°C & 2 minutes, 1 cycle of reverse transcription 53°C & 10 minutes, 1 cycle of activation 95°C & 2 minutes, and 40 cycles of amplification, including denaturation at 95°C for 03 seconds and extension 60°C for 30 seconds. Finally, results were interpreted using Applied Biosystems Interpretive Software, and Ct values for three target genes i.e., ORF1ab, N Protein, and S Protein of SARS-CoV-2 along with MS2 used as an internal control.

The samples were considered as positive if at least two of the primer probe sets showed amplification. The average Ct-value of a given sample was then converted to gene copy numbers considering the equivalence of 500 copies of SARS-CoV-2 genes as 26 Ct-value (provided with the kit), and the same was extrapolated to derive approximate copies of each gene. The average effective genome concentration present in a given sample was finally calculated by multiplying the RNA amount used as a template with each sample’s enrichment factor. Statistical Package for the Social Sciences (SPSS 21) has been used for hypothesis testing through Analysis of variance (ANOVA) and Duncan’s Multiple Range Test (DMRT). The OriginPro 2019b data analysis software has been used to draw boxplots.

## 3. Results and discussions

We detected and quantified variation in SARS-CoV-2 RNA from wastewater samples for three months (September and November) to understand the pandemic situation in Ahmedabad, Gujarat, India. Among the 116 samples analyzed in the study, 111 (95.7%) were found positive, comprising at least two positive RT-PCR results targeting SARS-CoV-2 ORF1ab, S gene, and N gene assays (**Table 1**). In addition to this, 109/116 (93.7%) samples showed positive RT-PCR results for each N, ORF 1b and S genes. The distribution analysis of Ct values for different genes using boxplot is represented in **Fig.2**. The average Ct values for N, ORF 1ab, and S genes were 32.50, 32.36, and 33.85, respectively. The average Ct values of internal control (MS2 bacteriophage) was 28.2, and no SARS-CoV-2 genes were detected in the negative control samples.

**Table 1.**
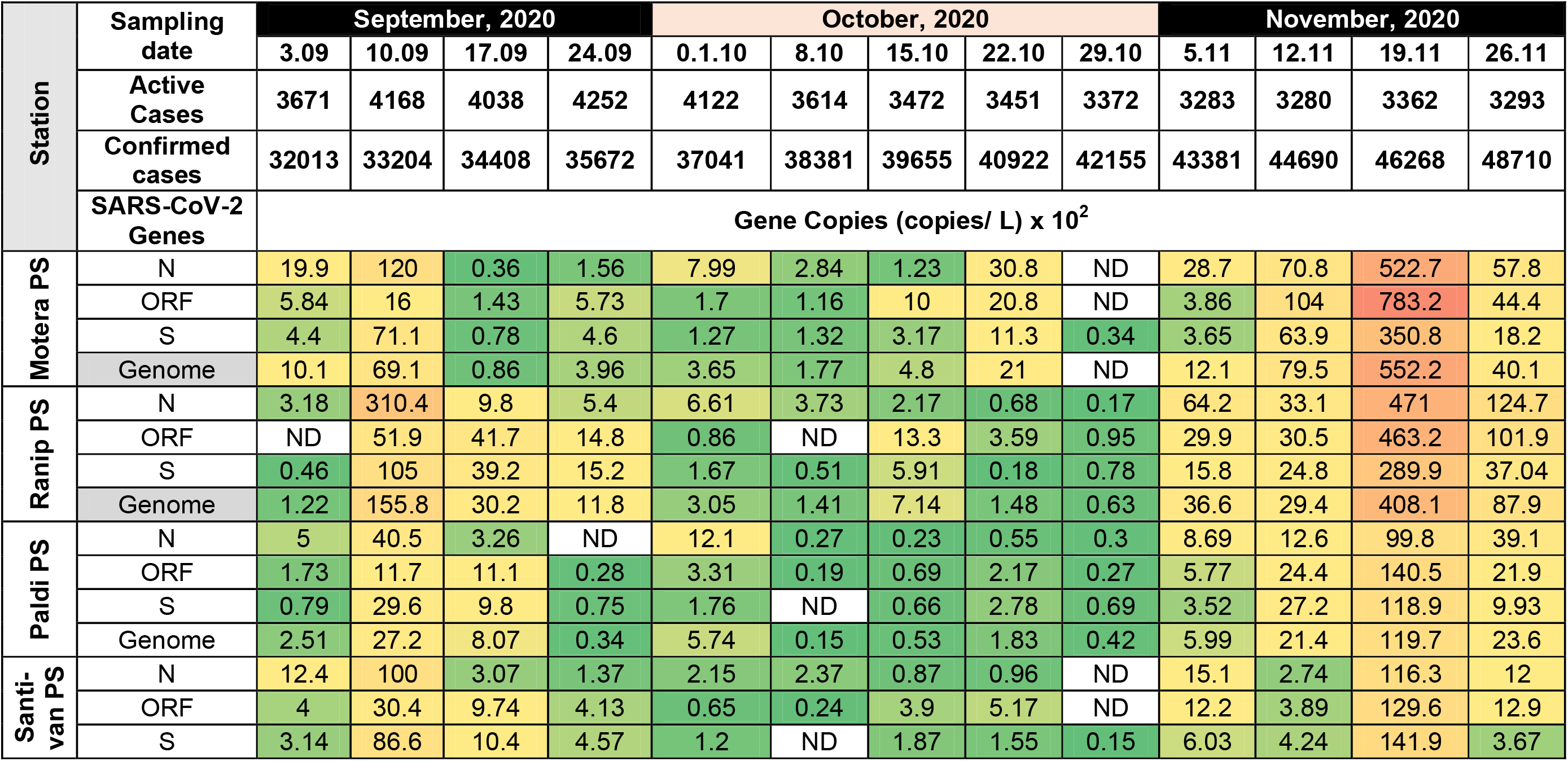
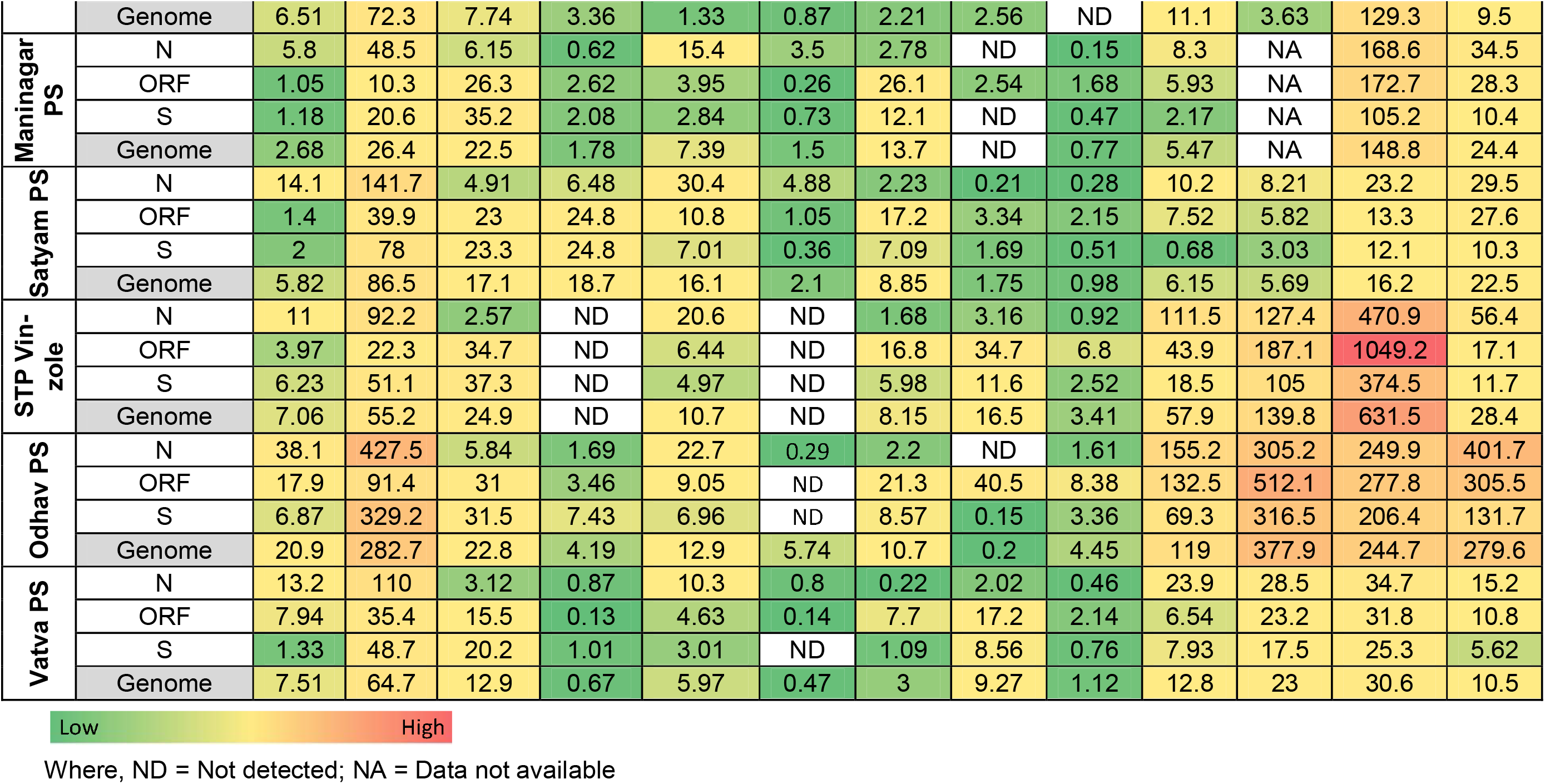
Temporal variation in gene copies of the SARS-CoV-2 targeted genes and genome concentration at various locations in Ahmedabad city

**Fig. 2.**
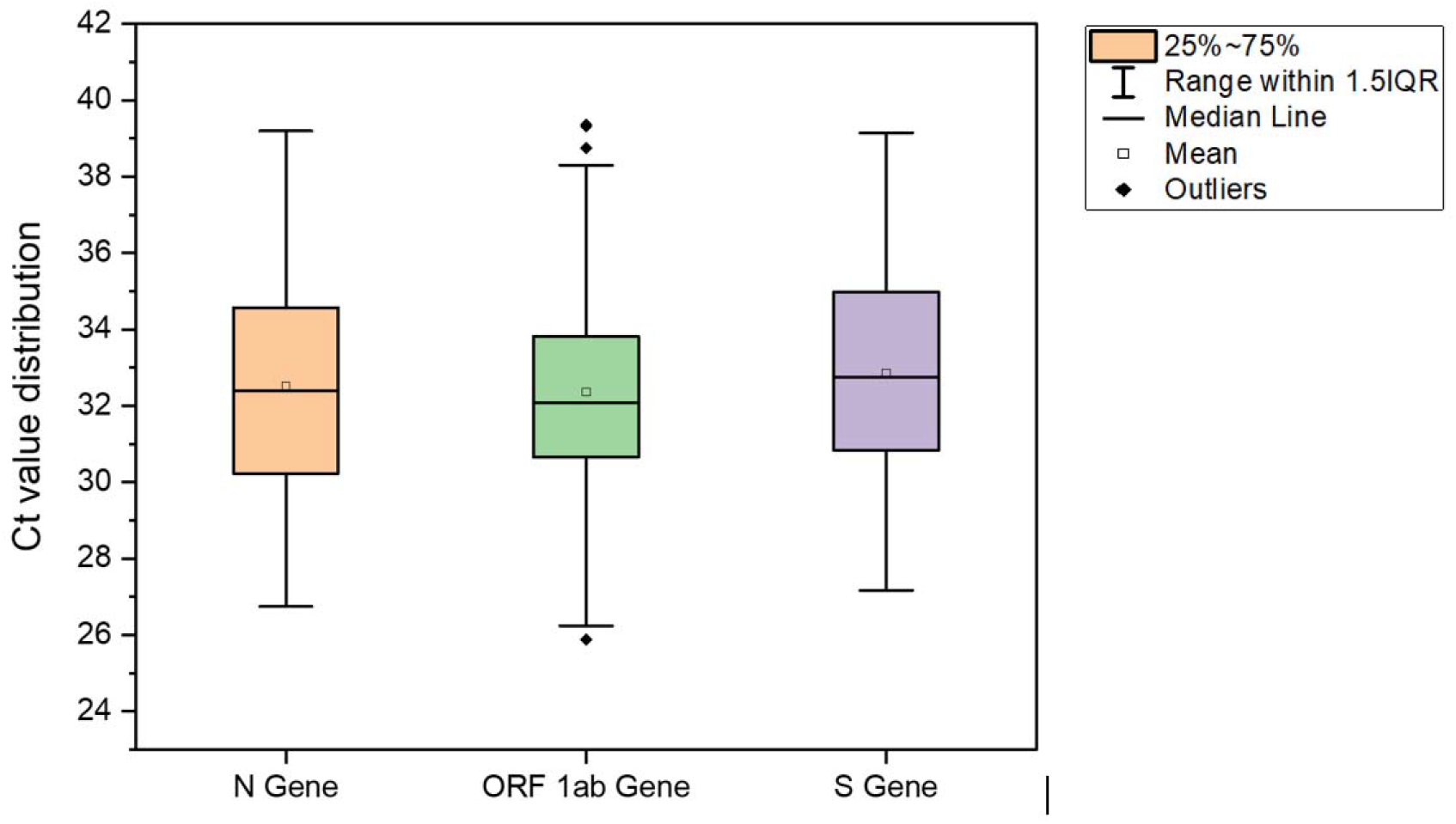
Distribution of Ct values of SARS-CoV-2 genes during the study period.

### 3.1 Monthly and Weekly Variations

Monthly variation depicted a significant decline of 89.7, 63.7, and 90.1% in N, ORF-1ab, and S gene concentration (copies/L), respectively in October compared to September 2020, followed by a sharp increment in November 2020 i.e. ∼25 folds in N gene, ∼22 folds in ORF 1ab and ∼26 folds in S gene. The PCR products for all three genes were maximum in wastewater samples of November, followed by September and October (**Fig. 3 a-c**). Likewise, the genome concentration of SARS-CoV-2 RNA was maximum in the month of November (∼10729 copies/ L), followed by September (∼3047 copies/ L), and October (454 copies/ L) in line with a ∼ 1.5-fold rise in the number of confirmed cases during the study period (3^rd^ September 2020 and 26^th^ November 2020) (**Fig. 3d**). Trends of monthly variation in SARS-CoV-2 RNA concentration in the wastewater samples may be ascribed to a decline of 19.3% in active cases in October 2020 and a rise of 1.82% in November 2020 compared to the preceding months. A little percentage increase of 1.82% in the active cases equaled 59 cases, while the total number of active cases was 3293 in the month of November 2020. However, at the same time, a prominent rise of 17.3% (i.e. 7386 new cases) noticed in November 2020. Also, a monthly decrease of 3.73% in recovered cases was noticed in November compared to October 2020. The monthly recovery rate of patients was 16.61, 19.31, and 15.58% in September, October, and November 2020, respectively. Apart from that, people’s casual and reluctant attitude during the festive season in India (mid-October to mid-Nov) might be the reason for the piercing rise in COVID-19 cases.

**Fig. 3.**
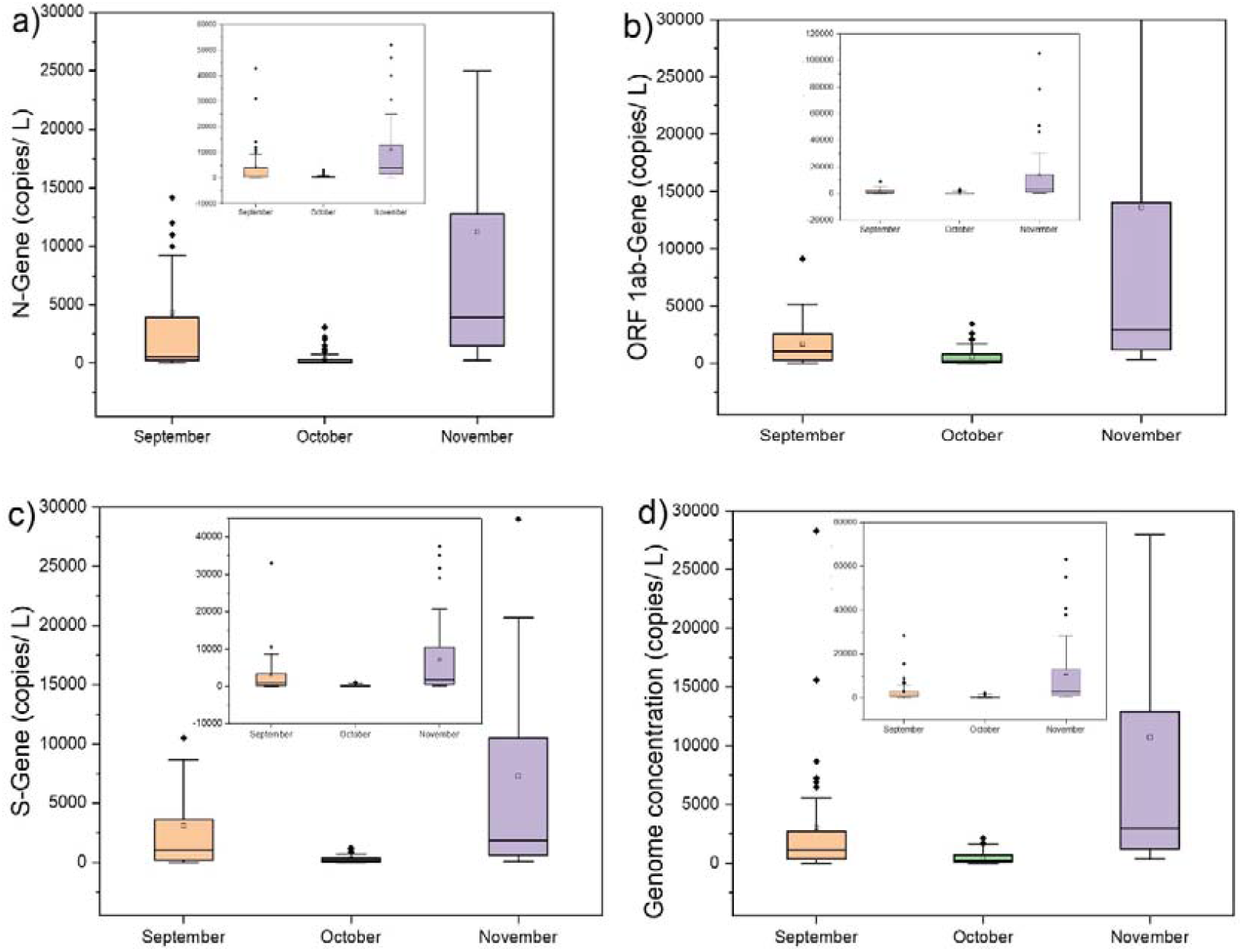
Distribution of SARS-CoV-2 gene copies on a temporal scale (monthly variation).

Weekly temporal variations in average SARS-CoV-2 gene copies were analyzed for SARS-CoV-2 RNA presence in samples collected from all the sampling locations in Ahmedabad and are displayed in **Fig. 4a-d**. One-way ANOVA and Duncan post hoc test (**p < 0.05**) were performed to see the significance level in gene copy variation among different sampling dates. The results showed significant differences in all three gene copies, i.e. N-gene (ANOVA, F= 7.49, p <0.001), ORF-1ab genes (ANOVA, F= 5.94, p <0.001), and S-gene (ANOVA, F= 8.25, p <0.001) on the temporal scale (sampling dates). Similarly, differences were significant in the case of genome concentration (ANOVA, F= 7.12, p <0.001). All three gene copies (i.e. N, ORF1ab, and S genes) and genome concentration were detected maximum on November 19th, 2020, and values were significant (p <0.05) as compared to other sampling dates. The exponential rise in virus geneconcentration might be due to the decline in the decreasing trend (< −0.1%, November 12^th^, 2020) followed by the increase in the number of active cases (i.e., 2.5% which corresponded to the 82 new cases on November 19^th^, 2020), compared to the earlier sampling dates.

**Fig. 4.**
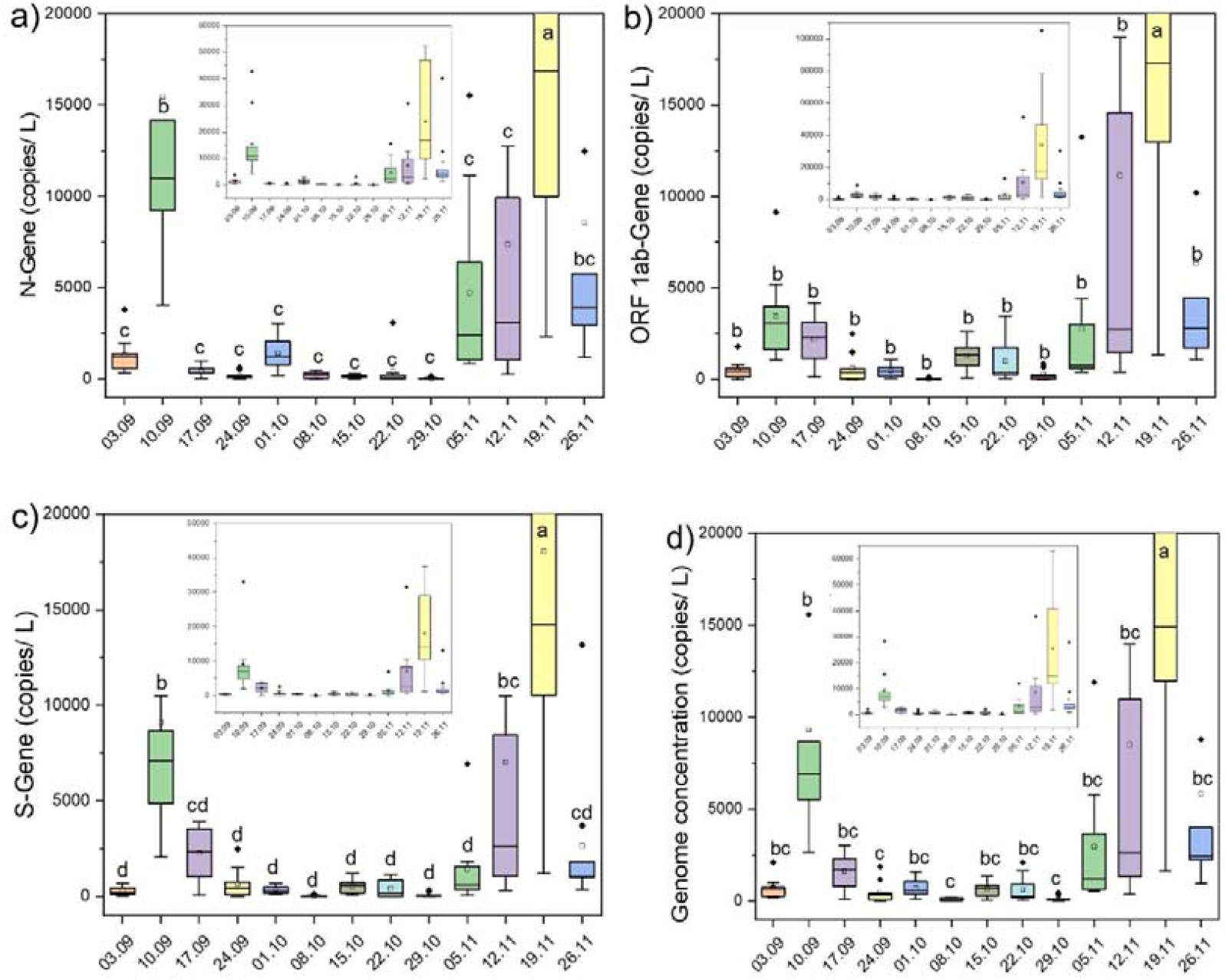
Temporal variations in targeted gene copies of SARS-CoV-2, collected from different sampling points a.) N gene, b.) ORF 1ab gene, c.) S gene, and d) Genome concentration

### 3.2 SWEEP-based city zonation and Identification of Hot-Spots

The SWEEP technology offers a better picture of the pandemic situation at the sub-city or zone level, relying on the SARS-CoV-2 RNA concentration in wastewater samples of a particular area. SWEEP data can help to estimate the actual extent of the infection due to the SARS-CoV-2, as it covers both asymptomatic and presymptomatic patients, which may be underestimated by clinical surveillance. Therefore, it would help in SWEEP data-based zonation of the city, and identifying hot-spots, which require more attention and rapid testing. On the other hand, clinical surveillance usually fails to classify the city into distinct zones based on the severity of the pandemic situation as it entirely depends on the data provided by the clinical centres that facilitate patients from the entire city, and dedicated manpower is required to maintain the demographic data. Also, sometimes-clinical survey-based secondary data is unreliable because it doesn’t include asymptomatic patients, presymptomatic patients, and patients who don’t go through clinical tests.

Depending on the genome concentration in wastewater samples based on analytical results, we identified highly susceptible areas for COVD-19 infection and its transmission among the community. The north (Motera and Ranip) and east (Odhav and Satyam) zones were highly affected areas with an average genome concentration of ∼15,574 and ∼13,397 copies/L, respectively, in November (**Fig. 5a**). Likewise, in September, wastewater samples collected from the east zone showed maximum genome concentration (∼5734 copies/ L), followed by the north zone (∼3536 copies/ L). Though areas present in north and east zones showed high virus genetic load, yet a sharp rise in SARS-CoV-2 RNA was noticed in all the zones in November 2020 (**Fig. 5a**). It has also been represented in a summarised format with comparison to the affected population in the city (**Fig.5b & c**). Overall, proper scrutiny and regular monitoring of wastewater could be useful for preparedness against adverse conditions as appeared in post-festive days in Ahmedabad.

**Fig. 5.**
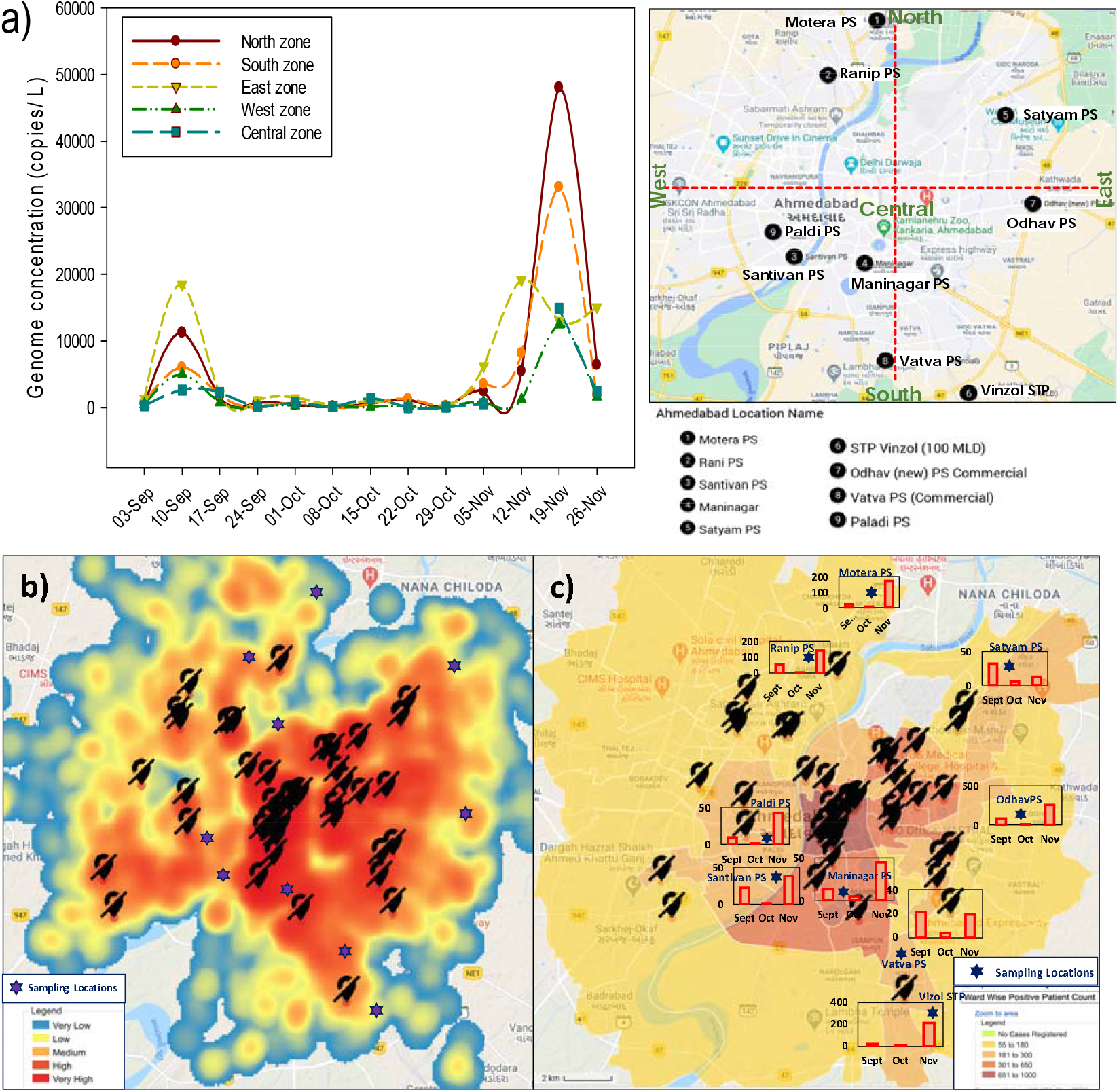
a) Zone-wise Covid-19 pandemic status in Ahmedabad city; b) Heat map of the overall infected population in Ahmedabad City obtained from http://google.org/crisismap/a/gmail.com/amdcovid19; c) Monthwise Effective genome concentration at the sampling locations (the present study) overlaying at wardwise positive patient count (http://google.org/crisismap/a/gmail.com/amdcovid19).

There are some studies available around the globe on early detection of SARS-CoV-2 RNA in wastewater, even before the first report of clinical diagnosis. For example, Madema et al. (2020) reported the presence of SARS-CoV-2 genetic material in wastewater in February, even before the official declaration of the first case in the Netherlands. Likewise, La Rosa et al. (2020) reported SARS-CoV-2 genetic material in wastewater samples before the first official documented report from two different cities in Italy. Similarly, Randazzo et al. (2020) detected SARS-CoV-2 RNA in wastewater samples from Spain. Since then, many researchers detected and reported the occurrence of SARS-CoV-2 RNA in wastewater samples and pondered its applicability for WBE surveillance (Ahmed et al., 2020, Kumar et al. 2020a, c). However, a few studies available focused on assessing its potential on the temporal scale in relation to the changes in COVID cases.

### 3.3 Early Warning Potential of WBE

In this view, the present research work followed our first proof concept study, where we detected SARS-CoV-2 genetic material in wastewater and proposed its wide applicability for COVID surveillance in the community (Kumar et al. 2020a). Examining the potential of WBE for COVID-19 surveillance as a potential tool showed that the percentage change in genome concentration level on a particular date was in conjunction with the confirmed cases registered 1-2 weeks later on a temporal scale by the regulatory authority based on clinical tests (**Fig. 6**). For example, on October, 8^th^, 2020, a sharp decline of ∼86% was noticed in the percentage change in the average genome concentration which was followed by ∼0.4% decline in the percentage change in confirmed COVID cases on October, 22^th^, 2020. Likewise, on November 5^th^, 2020, a steep hike of >22^nd^-folds in the percentage change in the average genome concentration was noticed compared to the earlier sampling date, which was followed by ∼0.6% and 2.37% increment in the percentage change in confirmed COVID cases on November 19^th^ and November, 26^th^, 2020, respectively. Therefore, we can predict the severity of the pandemic situation 1-2 weeks prior to the official reports by the regulatory body based on clinical tests.

**Fig. 6.**
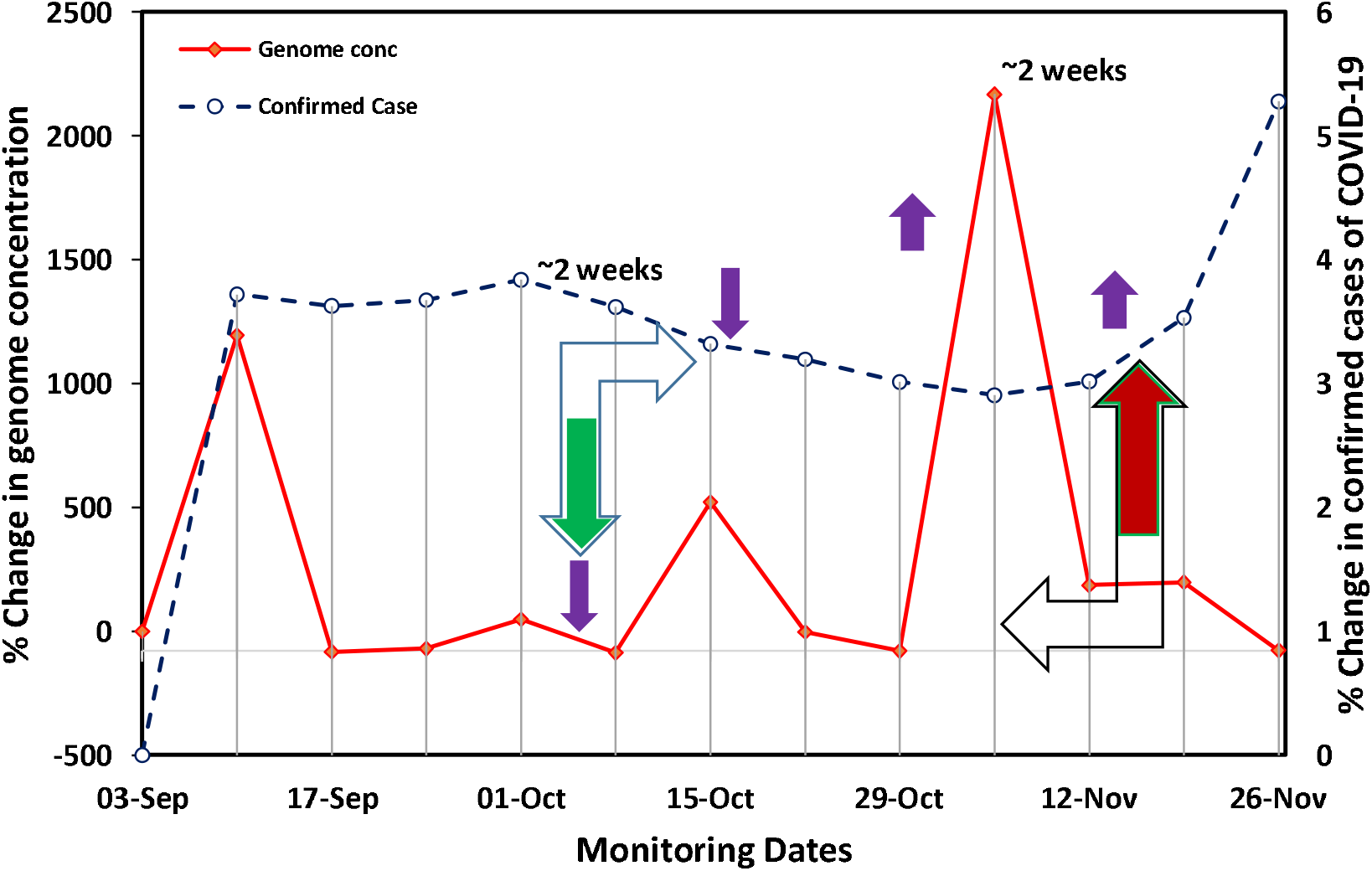
Potential and evidence of wastewater based epidemiology surveillance of Covid-19 pandemic as an early warning tool.

The results unravel the potential of WBE surveillance of COVID-19 as an early warning tool displayed by the adequate presence of SARS-CoV-2 genetic material in wastewater samples though limited cases were documented and based on the immediate future trends. These findings were in agreement with those of Ahmed et al. (2020b), who noticed a longitudinal decline in the presence of SARS-CoV-2 RNA with the tapering of the first epidemic wave; however, there was no concrete relationship between virus RNA and daily cases numbers.

### 3.4 Environmental Perspectives

Wastewater based epidemiology (WBE) has been proved highly effective in the early warning of COVID-19, however owing to its newness, there remains several aspects to be explored through continuous monitoring. The future work may focus on: i) To monitor the COVID-19 curve in the post-vaccination period through quantifying the genetic material of SARS-CoV-2 in the wastewaters of a given city (Ahmedabad); ii) To understand the association of antibiotic resistance with COVID-19 prevalence; iii) To develop an online portal with weekly update of genome concentration with accessibility provided to the public and policy makers; iii) To estimate the potential risk of SARS-CoV-2 in natural water bodies through various water activities using a quantitative microbial risk assessment (QMRA) framework; iv) To have a longer valuable time-series data to check the robustness of early warning capability of the techniques, and its possible benefits of making it accessible for public; and v) To develop predictive modelling to connect the missing points and for the advancement of SWEEP and support to other surveillance protocols.

Longer time-series data of SWEEP can be used for various modelling and risk evaluation study. It can also provide an additional way to understand the efficacy of vaccine through high resolution signs indicative of temporal variation in SARS-CoV-2 RNA. SWEEP can be taken into account for developing advisory in the context of rapid-testing, number of testing, community clearance, hotspot identification, vaccine need identification zones as well as, to stay at home the accurate scale of the pandemic must use the environmental surveillances of SARS-CoV-2 in wastewater to supplement the individual testing and timely identification.

In the first phase, we have explicitly shown the capability of WBE as early warning and city zonation tool. However, in a country like India, where sewer systems are not complete and treatment systems are not well-managed, it is important to have long-term monitoring for a year at the least so that precious meaningful data for the developing country can be obtained. Furthermore, a practical guide and pandemic management tools can be developed by integrating the virtues of information technology with early warning capability of waste-water surveillance. A confidence may be generated by inculcating informed understanding among the commons about the effectiveness of treatment plants through environmental surveillance of COVID-19, detecting the genetic material of SARS-CoV-2 in sewage. So that the government agencies like AMC can be convinced to incorporate WBE for any COVID-like epidemic/pandemic outbreak in the future.

## 4. Conclusion

A temporal variation of SARS-CoV-2 RNA presence in wastewater was studied for a period of three months in Ahmedabad, India. A total of 111 samples (95.7%) of the total 116 samples tested in the study were found to be positive, with at least two positive RT-PCR results targeting SARS-CoV-2 ORF1ab, S gene, and N gene assays. Monthly variation depicted a significant decline in all three gene copies in October compared to September 2020, followed by a sharp increment in November 2020. Correspondingly, the descending order of average genome concentration was November (∼10729 copies/ L) > September (∼3046 copies/ L) > October (∼453 copies/ L). This finding was further supported by the relation between the percentage change in genome concentration level and confirmed cases, which followed a similar trend on the temporal scale with a ∼1 to 2 week’s time distance. Also, the genome concentrations at various sampling locations were well correlated with the number of cases in a particular zone. The results unveiled the untapped potential of WBE surveillance of COVID-19 as an early warning tool for practical use of city zonation based on SWEEP data for actual scenarios and future prediction. This approach may help the authorities identify the hotspots within a city and tuning effective management interventions. Further research may be focused on quantifying the correlation of SWEEP results with clinical surveillance data and developing a predictive model that can translate SWEEP data for easy propagation to policymakers and the general public to enhance the preparedness and management of pandemics.

## Data Availability

All data are included in the paper. If anything else required we will provide it.

## Notes

The authors declare no competing financial interest.

## Acknowledgement

This work is funded by UNICEF, Gujarat and UKIERI. We also acknowledge the help received from Dr. Arbind K Patel, Mr. Alok Thakur, and other GBRC staffs who contributed towards sample and data analyses.

